# Investigating the relationship between climatic factors and malaria transmission dynamics in Southwest states of Nigeria

**DOI:** 10.1101/2024.05.09.24307136

**Authors:** Owolabi Yusuf, Chukwu Okoronkwo, Cyril Ademu, Sheetal P. Silal

## Abstract

**Background:** In order to combat malaria, numerous strategic initiatives have been deployed over the years, with the goal of eliminating the disease in Nigeria. Despite these efforts, malaria morbidity still persists across the nation, and is presently under control measures. Various factors, encompassing both climatic and non-climatic elements, significantly impact the dynamics of malaria transmission across the country. This study applied time series analysis methods to understand the relationship between climatic factors, particularly temperature and rainfall, and the occurrence of malaria in the Southwest area of Nigeria.

**Methods:** This study analysed monthly data on documented cases of uncomplicated malaria obtained from the District Health Information System version 2, overseen by the Federal Ministry of Health. Additionally, monthly rainfall and temperature data spanning from 2013 to 2021, acquired from the Nigeria Meteorological Agency, were analysed. The datasets used for analysis were accessed on June 7, 2022, for research purposes. We applied univariate time series models to analyse monthly data on average temperature, rainfall, and malaria cases. Subsequently, cross-correlation analyses were conducted to investigate the relationships between malaria case data and the corresponding rainfall and temperature patterns, following a pre-whitening process.

**Results:** The patterns of malaria transmission exhibit variability among the various states within the Southwest region of Nigeria from 2013 to 2021. On average, Oyo State consistently recorded relatively higher temperatures compared to its neighbouring states. Conversely, Ondo State receives a higher average monthly rainfall. This state, similar to others in the region, experiences two distinct wet seasons, each characterised by different peak periods.

In terms of temporal relationships, it is predicted that rainfall precedes malaria cases by one month across all the states. However, with regard to temperature, the five other states (excluding Osun state) lead by two months.

**Conclusion:** The results of this study underscore the varied characteristics of malaria transmission patterns and climate change, even across the individual states within the Southwest region of the country. This recognition of variations in malaria transmission and climatic factors is valuable for policymakers as it enables them to strategically allocate and optimise the implementation of interventions at state-specific level, rather than at zonal level. These interventions might involve distributing and advocating for the use of insecticide-treated bed nets, indoor residual spraying, and seasonal malaria chemoprevention tailored to the unique requirements and circumstances at the state level.

## Introduction

Malaria presents a considerable public health challenge on a global scale, as highlighted in the 2023 WHO report on malaria, this disease caused approximately 249 million cases and led to around 608,000 fatalities in a majority 85 countries (1). Alarmingly, the African region under the purview of the World Health Organization (WHO) faced an exceptionally disproportionate burden, representing 94% of reported malaria cases and 95% of malaria-related fatalities. Notably, approximately 80% of these fatalities occurred in children under the age of five (2).

It continues to be a significant health challenge in nations throughout Sub-Saharan Africa, causing considerable harm, especially in Nigeria, which reports the highest number of cases and deaths (2,3). Despite the implementation of measures by the National Malaria Elimination Programme (NMEP) and other stakeholders to combat this disease, malaria remains endemic across the country, and the risk of transmission exists year round (4,5). The seriousness of malaria in Nigeria can be attributed in part to the country’s climate and weather, which favour the growth of malaria parasites and the reproduction of the vectors. Conversely, temperate regions of the world experience minimal malaria occurrences, due to their unfavourable climatic conditions for parasite development (6,7). Consequently, malaria remains a pressing health challenge for developing countries, especially those located in the world’s tropical and subtropical regions (8).

The influence of climatic factors, including temperature and precipitation, on the dynamics of malaria transmission has been well-documented, extending to various African contexts (9–14). These factors are crucial in influencing the survival, growth, and behaviour of both the malaria parasite and the mosquito vectors that carry it and facilitate its transmission. Consequently, deciphering the intricacies of how climatic variables modulate malaria transmission dynamics proves a fundamental step in the fight against the disease in endemic regions.

The Southwest states of Nigeria, of Oyo, Osun, Ogun, Ekiti, Ondo, and Lagos, exhibit a diverse range of climatic patterns. These states experience variations in both temperature and precipitation levels, due to the presence of distinct wet and dry seasons. Such regional disparities in climate, combined with variations in ecological setting, have the potential to exert an influence on the patterns of malaria transmission. However, the scarcity of comprehensive studies that investigate the specific relationships between climatic factors and malaria dynamics within Southwest Nigeria underscore the need for further research.

As far as we know, this is the inaugural research endeavour of its nature investigating the relationship of meteorological factors such as rainfall and temperature on malaria morbidity across each state of the Southwest zone of Nigeria, applying time series analysis will be conducted using unified data from the District Health Information System version 2 (DHIS2). While some studies have examined malaria morbidity across the country (15,16), it is essential to recognise that addressing malaria effectively requires interventions tailored to specific contexts and the regional dynamics of transmission. In this regard, a single approach may not adequately account for the diverse factors influencing malaria prevalence and control strategies in a country that continues to record the peak cases and fatalities due to malaria (2).

By recognizing the differences in patterns and links between climatic factors and malaria occurrences across the Southwest states, data from DHIS2 can provide valuable insight, which can serve as a foundation for developing effective tools to assess and implement appropriate interventions to combat malaria.

A few studies have investigated the connections among rainfall, temperature, and malaria incidence at a smaller scale within local government areas, focusing on specific states in the country (17–19). This study seeks to explore the influence of rainfall and temperature on confirmed uncomplicated malaria cases across each state of the Southwest zone of Nigeria using time series analysis.

### Study area and data

The study area includes Nigeria’s Southwest zone, which constitutes one of the country’s six geopolitical regions. This zone encompasses six individual states, specifically Oyo, Osun, Ogun, Ekiti, Ondo, and Lagos (Fig 1). Geographically, this area is situated between longitudes 20°31’E and 6°00’E and latitudes 6°21’N and 8°37’N, covering a total land area of approximately 77,818 square kilometres (20).

**Fig 1.**
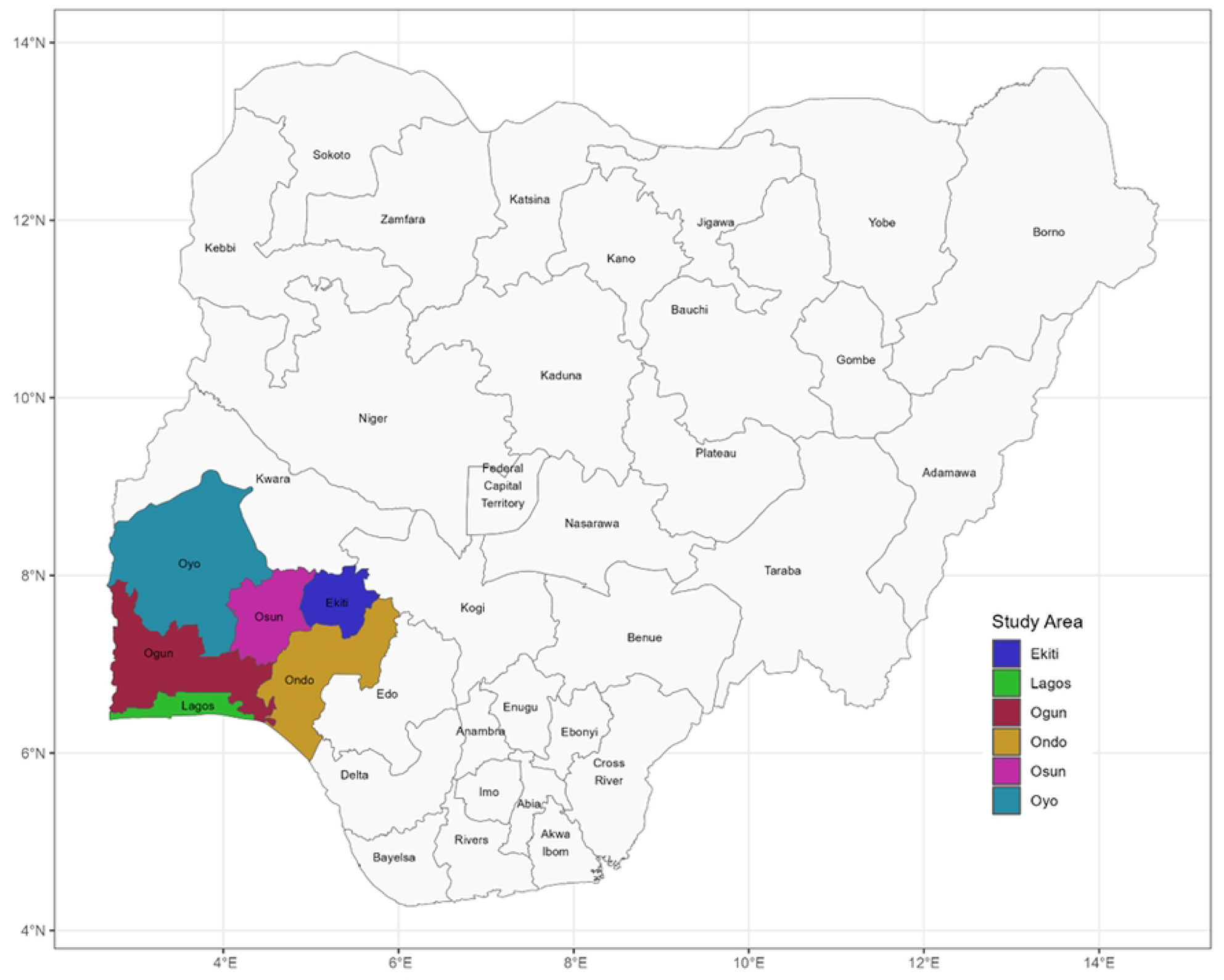
Visualisation of Nigeria Depicting the Southwestern Zone. (Data source: https://www.igismap.com/download-nigeria-shapefile-free-administrative-boundary-state-city-map/)

The climate in this area exhibits two clearly defined seasons: a rainy period occurring from April to October, followed by a dry season lasting from November to March. Temperature fluctuations within this zone typically range from 21 to 35 degrees Celsius (°C), accompanied by a relatively high humidity level of approximately 77% (21).

Monthly healthcare facility records of confirmed malaria cases across the six states in Southwest Nigeria for the period 2013-2021 were sourced from the NMEP. Monthly temperature data, including both minimum and maximum temperatures, along with rainfall data (measured in millimetres), were acquired from the Nigeria Meteorological Agency (NiMet) for the identical time frame as the malaria dataset.

This study’s research protocol obtained ethical clearance from the Research Ethics Committee of the Faculty of Science at the University of Cape Town. Furthermore, official permission and consent to utilize malaria data collected through the District Health Information System 2 (DHIS2) were obtained from the National Malaria Elimination Programme (NMEP) in Nigeria.

### Analytical methods/approach

In this study, we employed a time series analysis method to scrutinize the monthly trends in malaria cases. Initially, we disintegrated the time series data into its fundamental segments, which encompass the seasonal, trend, and residual components, employing the locally estimated scatterplot smoothing (Loess) technique. This disintegration facilitated a quantitative evaluation of both the trend and seasonality within the time series, depicted as y_t=T_t+S_t+R_t. In this expression, T_t signifies the smoothed trend component, S_t corresponds to the seasonal component, and R_t represents the residual component (22).

To investigate the connection between monthly confirmed uncomplicated malaria cases and climatic variables like rainfall and average temperature, we developed cross-correlation functions. Moreover, we utilized autocorrelation function (ACF) and partial autocorrelation function (PACF) to examine the autocorrelation within the stationary series. The aim here was to reveal any possible temporal relationships inherent in the dataset.

ARIMA models, which stand for Autoregressive Integrated Moving Average, were employed to model malaria incidence, rainfall, and temperature data for each state. The choice of the most appropriate ARIMA models was determined based on various criteria, such as the Akaike information criteria (AIC), the Box-Ljung test for assessing goodness of fit, and the scrutiny of normal Q-Q plots of residuals. The objective was to verify that the selected models adequately represented the inherent patterns and variations in the data.

To investigate the specific time lags at which the rainfall and temperature series correlated with malaria cases, cross-correlation functions (CCF) were used following the pre-whitening of the series. This procedure helped us identify significant lag terms and potential correlations between climatic factors and malaria transmission.

The R version 2021.09.0+351 Copyright (C) 2009-2021 were used for the data preparation and analyses. The datasets used for analysis were accessed on June 7, 2022, for research purposes and analyses were conducted individually for each of the six states in the Southwest region, namely Oyo, Osun, Ogun, Ekiti, Ondo, and Lagos.

## Results

### Trends in uncomplicated malaria cases, rainfall, and temperature patterns

Figs 2 and 3 depict the case patterns for six states, along with average temperature and rainfall data spanning from 2013 to 2021. While an initial observation of these figures does not readily reveal the seasonal patterns in malaria morbidity, Fig S3 offers a more detailed view of these patterns. These figures demonstrate a consistent upward trend in malaria cases across all six states during the specified period.

**Fig 2.**
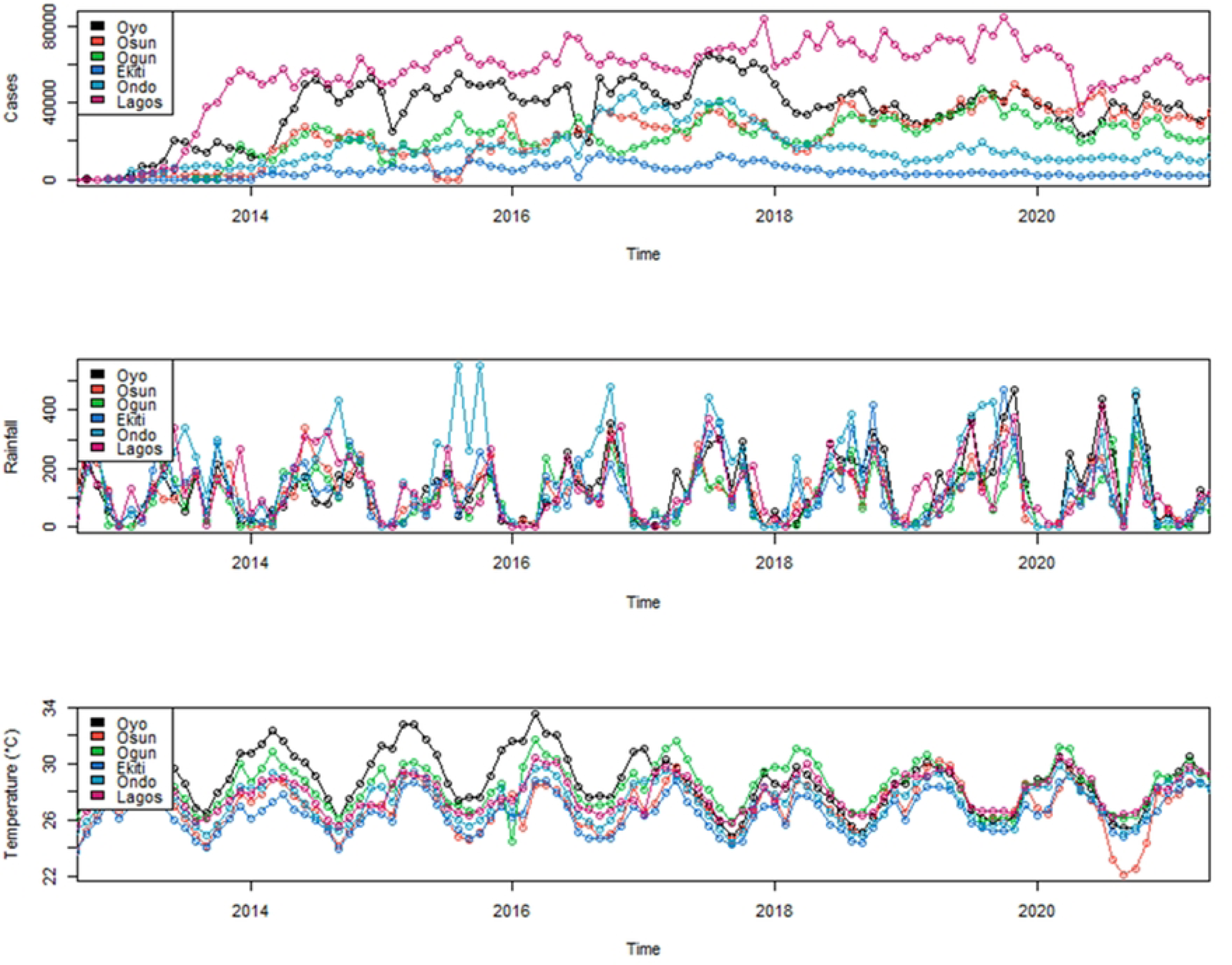
Displays the annual trends in uncomplicated malaria cases, average rainfall (mm), and average temperature (°C) across the six Southwest states.

**Fig 3.**
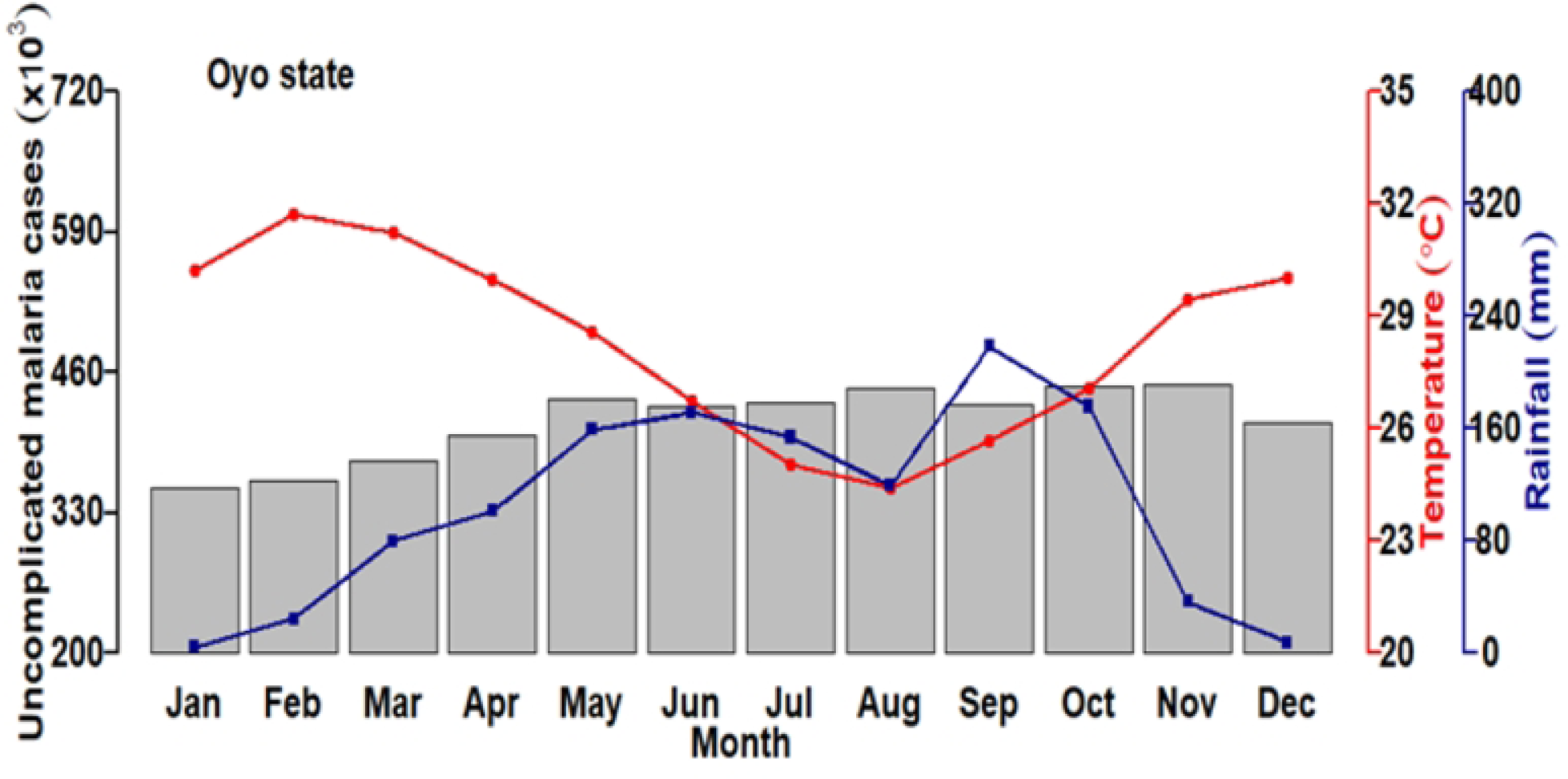
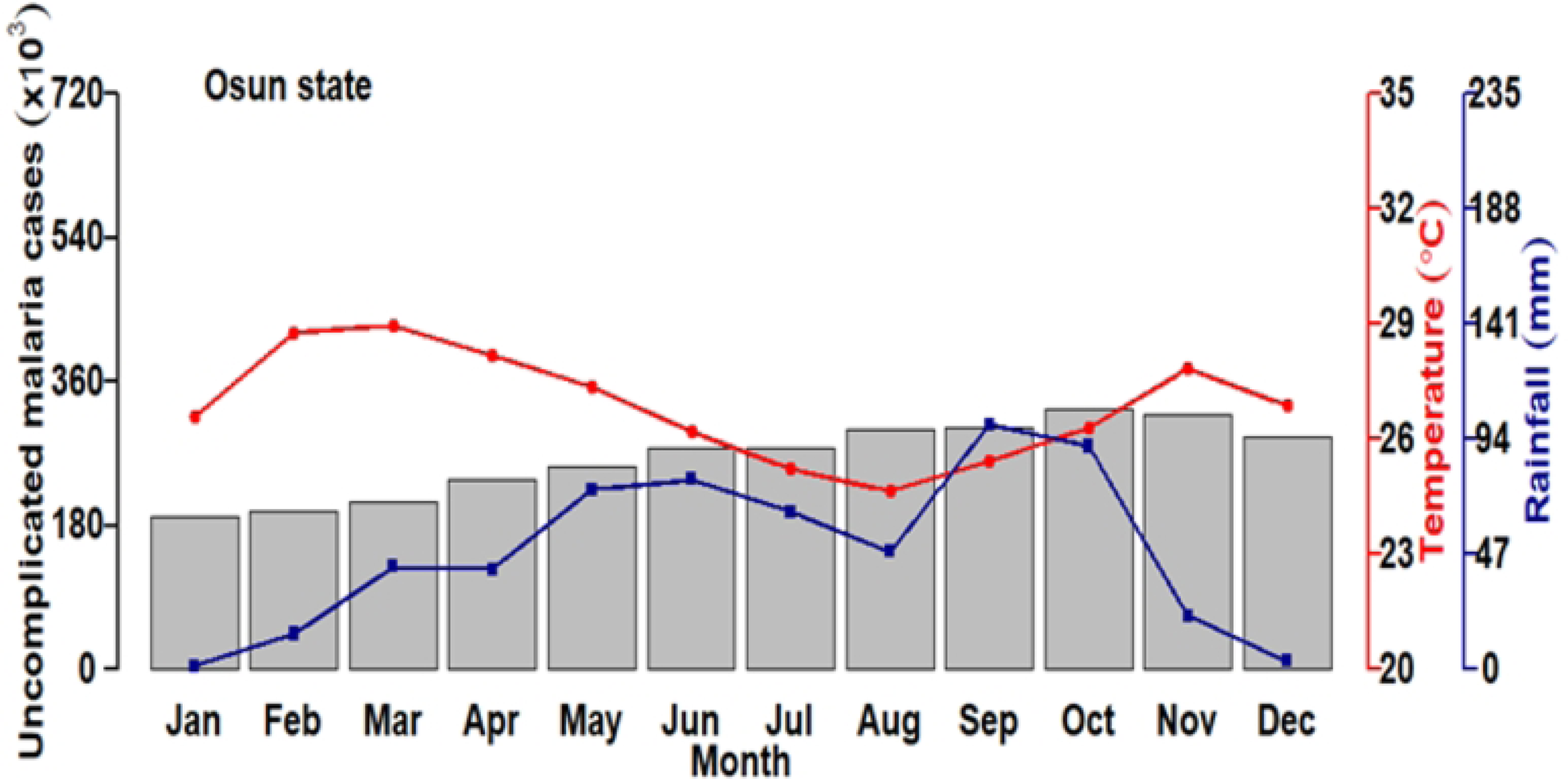
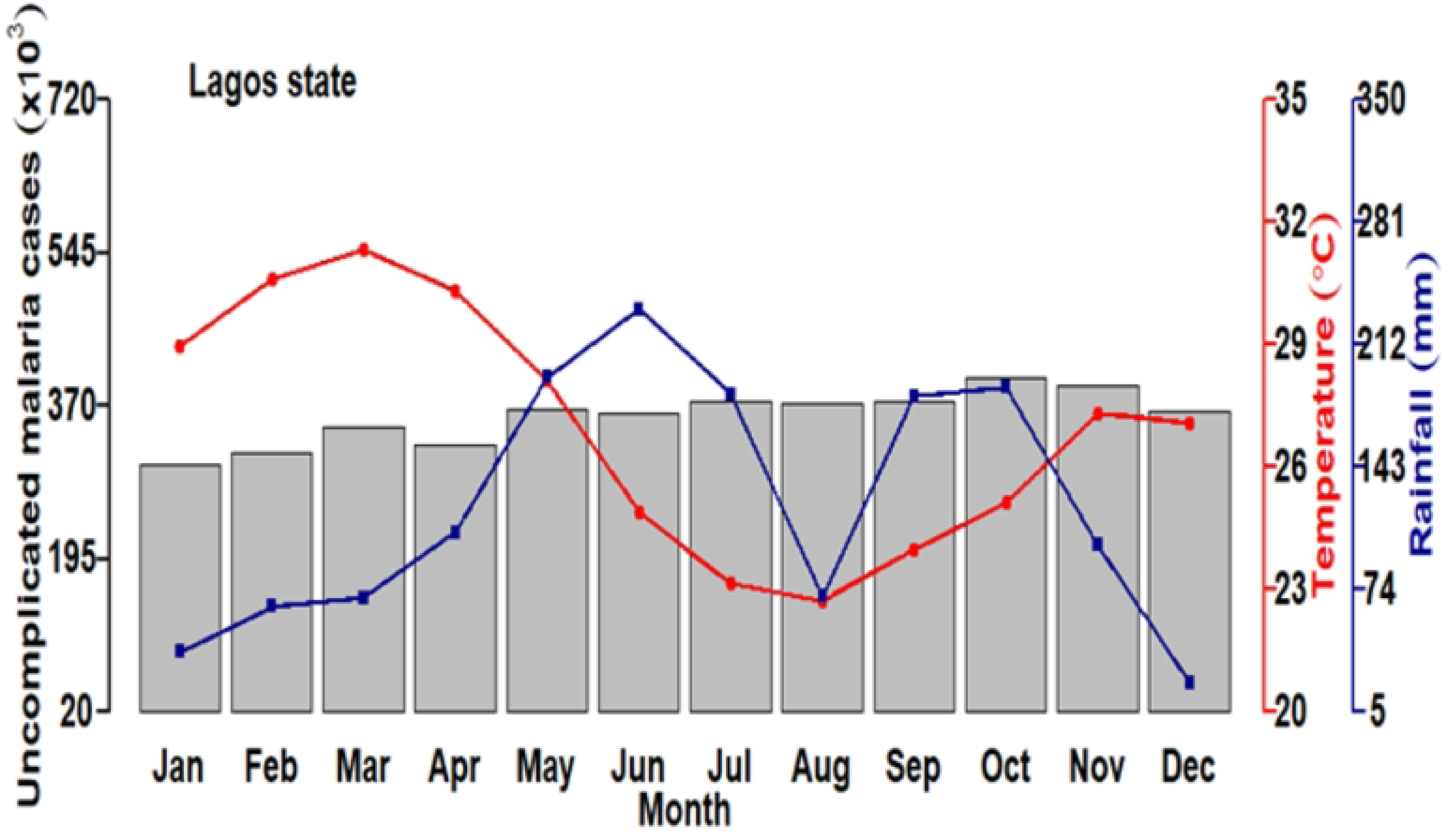
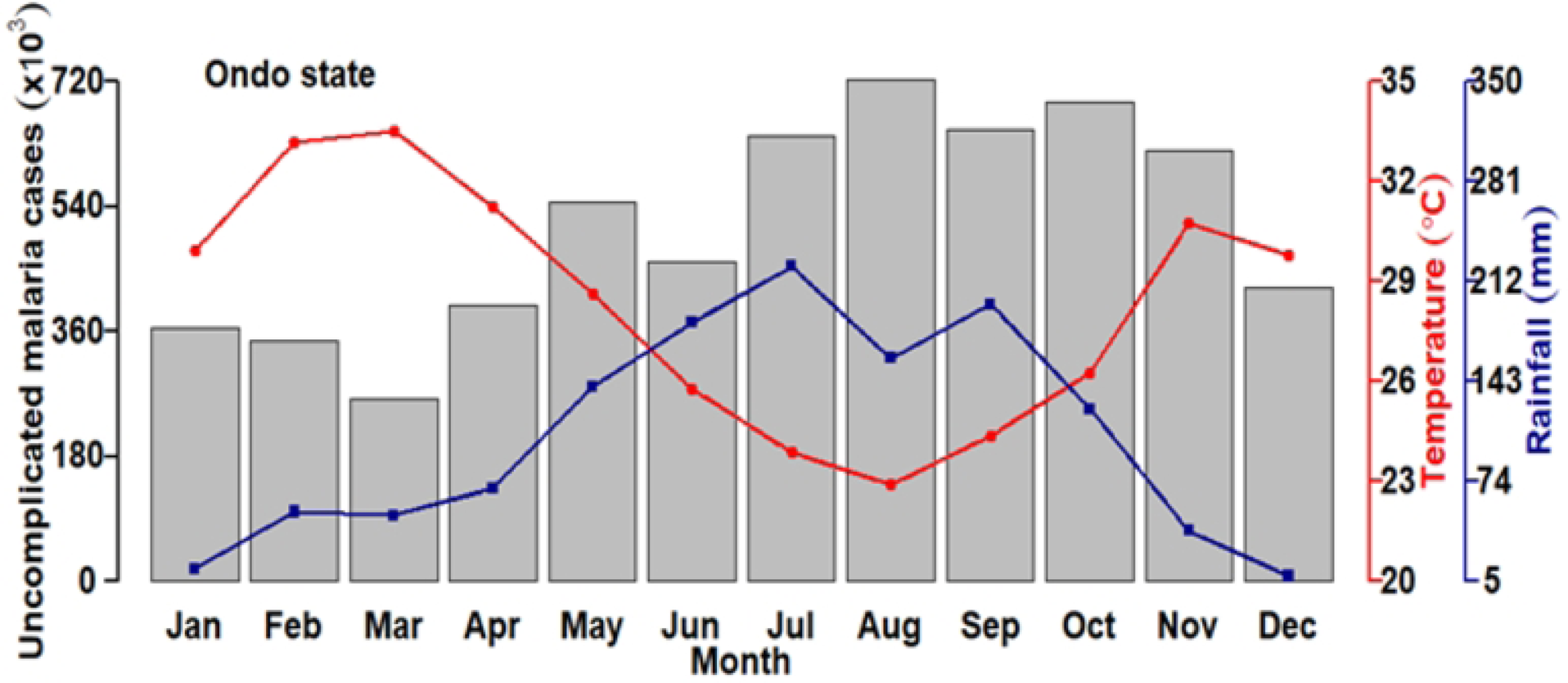
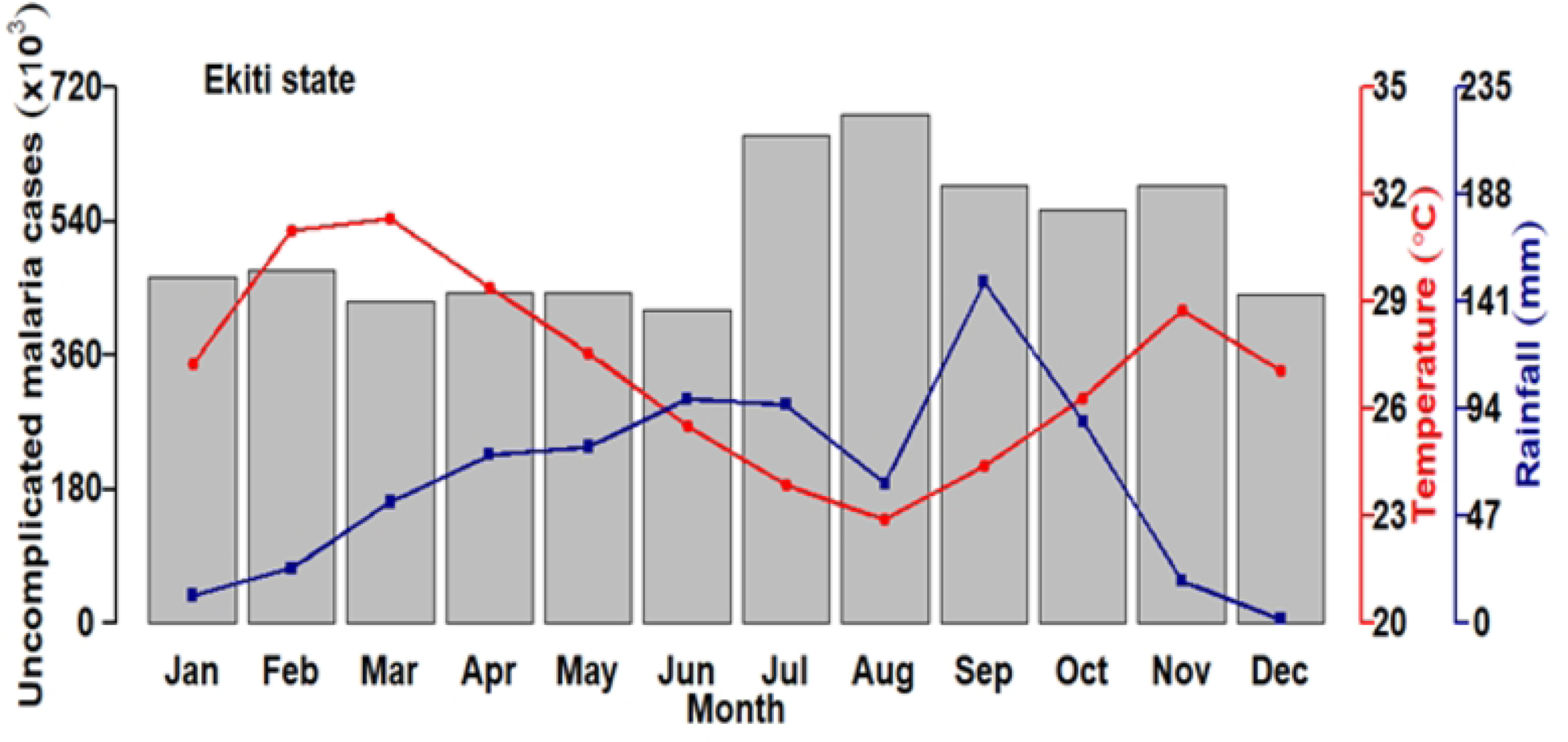
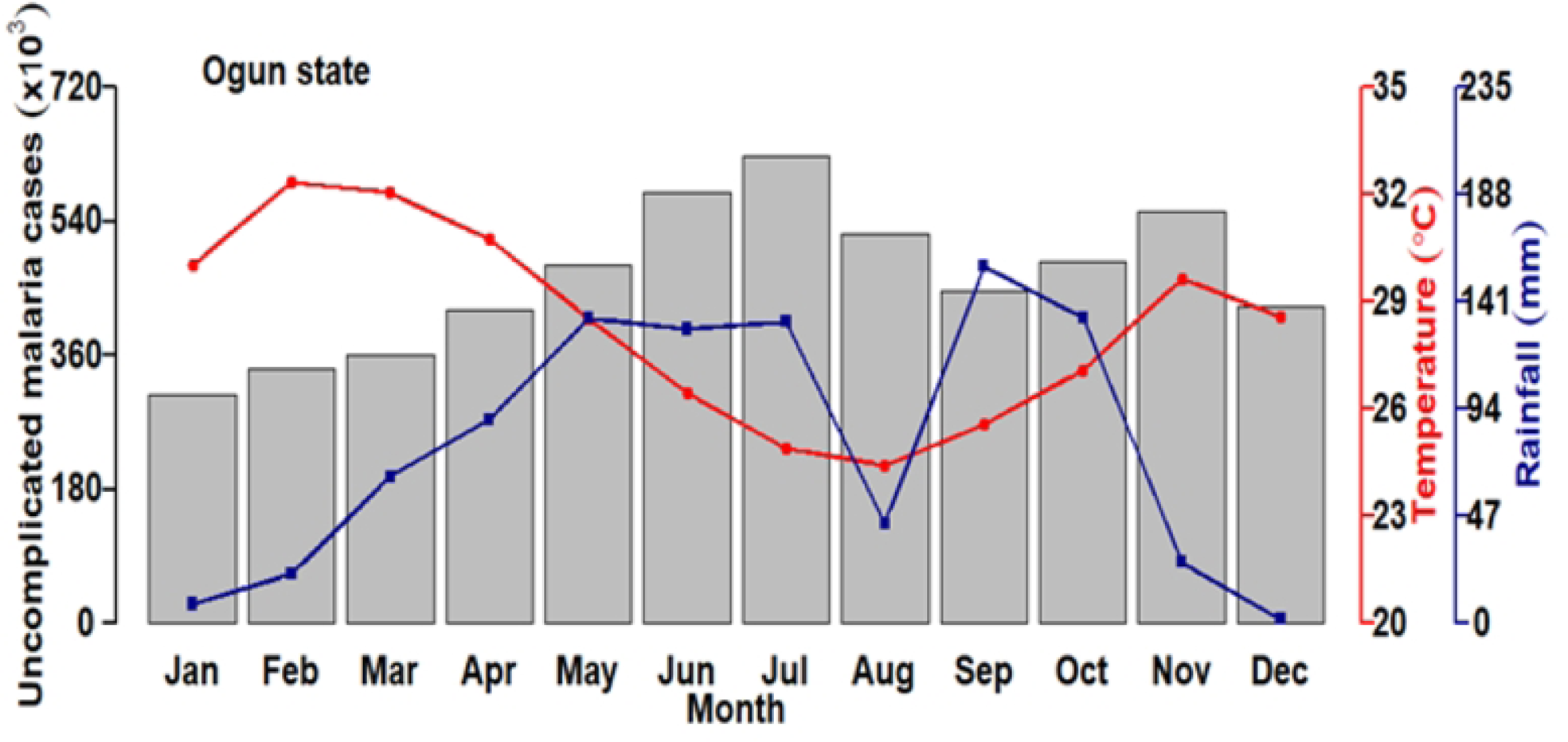
Patterns for aggregated uncomplicated malaria cases for the six states (a) Oyo, (b) Osun, (c) Ogun, (d) Ekiti, (e) Ondo, (f) Lagos by month for 2013 to 2021.

During the years 2013 to 2021, Oyo state reported an average monthly malaria case count of 38,883. Monthly cases ranged from a minimum of 2,898 to a maximum of 65,283. The monthly average incidence of uncomplicated malaria in this state equated to 5.17 cases per 1000 individuals, a calculation based on the estimated population of 7,512,855 as of 2019 (sourced from the National Bureau of Statistics, Nigeria).

Precipitation in the state averaged 134.78 mm monthly, with the lowest recorded rainfall at 0.0 mm and the highest at 470.10 mm. Furthermore, in Oyo state, the mean monthly temperature stood at 28.77°C, with the lowest and highest average temperatures recorded at 24.81°C and 33.56°C, respectively (as outlined in Table 1).

**Table 1.** Summary uncomplicated malaria for the six Southwest states of Nigeria from 2013-2021.

Persons with uncomplicated malaria (n)
|  | Oyo | Osun | Ogun | Ekiti | Ondo | Lagos |
| --- | --- | --- | --- | --- | --- | --- |
| Minimum | 2898 | 0 | 0 | 0 | 2676 | 66 |
| Median | 40315 | 28045 | 24332 | 3222 | 14112 | 61635 |
| Mean | 38883 | 25612 | 23162 | 4258 | 16659 | 58750 |
| Maximum | 65283 | 49904 | 47861 | 13511 | 44862 | 84691 |

**Table 2.** Summary rainfall for the six Southwest states of Nigeria from 2013-2021.

|  | Oyo | Osun | Ogun | Ekiti | Ondo | Lagos |
| --- | --- | --- | --- | --- | --- | --- |
| Minimum | 0.00 | 0.00 | 0.00 | 0.00 | 0.00 | 0.00 |
| Median | 122.35 | 98.05 | 88.60 | 107.10 | 126.05 | 112.25 |
| Mean | 134.78 | 114.73 | 102.59 | 124.88 | 158.80 | 136.84 |
| Maximum | 470.10 | 341.30 | 313.10 | 469.30 | 552.40 | 413.80 |

Interestingly, there seems to be an inverse relationship between monthly rainfall and malaria incidence in Oyo State. Malaria incidence rises from January, peaking in August, which corresponds to the lowest peak in temperature and a slightly lower peak in rainfall. Subsequently, there is a slight decline in malaria incidence, coinciding with the peak of rainfall and a rise in temperature.

In Osun State, the average monthly malaria cases recorded between 2013 and 2021 totaled 25,612, resulting in an average monthly incidence rate of 6.04 cases per 1000 individuals as presented in Table 4, calculated using the 2019 projected population of 4,237,396, as per the National Bureau of Statistics, Nigeria. Malaria cases in Osun state varied significantly, with a minimum of 0 cases (indicating months without reported cases) and a maximum of 49,904 cases during this period.

In terms of rainfall, Osun state recorded an average of 114.73 mm, with the lowest of 0.00 mm and highest of 341.30 mm during the same period. Fig 3 illustrates the patterns of uncomplicated malaria in Osun State, showcasing two distinct peaks in malaria incidence closely mirroring the rainfall patterns seen in Fig 2 in the middle panel.

The increase in malaria instances usually begins a few months following the start of the rainy season, with the highest malaria occurrence closely corresponding to the peak rainfall during both the initial and second wet seasons, as illustrated in Figure 3.

Ogun State exhibited a distinctive bimodal pattern of malaria incidence, as indicated in both Fig 2 and 3. From 2013 to 2021, an average of 23,162 malaria cases were reported monthly, resulting in an average monthly incidence rate of 3.89 cases per 1000 individuals, calculated using the 2019 projected population of 7,512,855 from the National Bureau of Statistics, Nigeria. The average, lowest, and highest values for rainfall and temperature during this period in Ogun State were 102.59 mm, 0.00 mm, and 313.10 mm, and 28.36°C, 24.42°C, and 31.70°C, respectively, as presented in Table 1.

Ekiti State displayed two pronounced rainfall peaks in June and September, closely matching the observed patterns of malaria cases in September, as illustrated in both Fig 2 and 3. Over the period from 2013 to 2021, an average of 4,258 malaria cases were reported monthly, resulting in an average monthly incidence rate of 1.27 cases per 1000 individuals, calculated using the 2019 projected population of 3,350,401 from the National Bureau of Statistics, Nigeria. Malaria cases in Ekiti State ranged from 0 (representing months without any reported cases) to 13,511 cases.

Additionally, the average, lowest, and highest values for rainfall in Ekiti State during this period were 124.88 mm, 0.00 mm, and 469.30 mm, respectively. The average monthly temperature in Ekiti State was 26.47°C, with lowest and highest temperatures of 23.88°C and 29.14°C, respectively, as indicated in Table 1.

In Ondo State, two distinct rainfall peaks occurred in July and September, with the highest number of malaria cases observed in August, as shown in Fig 3. From 2013 to 2021, an average of 16,659 malaria cases were reported monthly, resulting in an average monthly incidence rate of 3.35 cases per 1000 people. These calculations were based on the 2019 projected population of 4,969,707 provided by the National Bureau of Statistics, Nigeria. Malaria cases in Ondo State ranged from 2,676 to 44,862 during this period.

Furthermore, the average, lowest, and highest values for rainfall recorded in Ondo State during this period were 158.80 mm, 0.00 mm, and 552.40 mm, respectively. The average monthly temperature in Ondo State was 27.35°C, with lowest and highest temperatures of 24.40°C and 29.85°C, respectively, as indicated in Table 1.

Lagos State encountered two significant spikes in rainfall, particularly in June and September. The peak occurrence of malaria cases was observed in October, as shown in Figure 3. The coastal location of Lagos may contribute to this pattern. Over the period from 2013 to 2021, an average of 58,750 malaria cases were reported monthly, and the average monthly incidence rate was 4.59 cases per 1000 people, calculated based on the 2019 projected population of 12,772,884 from the National Bureau of Statistics, Nigeria. Malaria cases in Lagos State ranged from 66 to 84,691 during this time frame.

Additionally, the average, minimum, and maximum amounts of rainfall in Lagos State during this period were recorded as 136.84 mm, 0.00 mm, and 413.80 mm, respectively. The mean monthly temperature in Lagos State stood at 27.89°C, with the lowest and highest temperatures values as 25.49°C and 30.43°C, respectively, as illustrated in Table 3.

**Table 3.** Summary Temperature (°C) for the six Southwest states of Nigeria from 2013-2021.

|  | Oyo | Osun | Ogun | Ekiti | Ondo | Lagos |
| --- | --- | --- | --- | --- | --- | --- |
| Minimum | 24.81 | 22.09 | 24.42 | 23.88 | 24.40 | 25.49 |
| Median | 28.80 | 27.07 | 28.49 | 26.41 | 27.64 | 27.95 |
| Mean | 28.77 | 26.87 | 28.36 | 26.47 | 27.35 | 27.89 |
| Maximum | 33.56 | 30.20 | 31.70 | 29.14 | 29.85 | 30.43 |

**Table 4.** Malaria incidence per 1000 population by state (2013-2021).

| State | Average Monthly Malaria Cases | Average Monthly Incidence (per 1000) |
| --- | --- | --- |
| Oyo | 38,883 | 5.17 |
| Osun | 25,612 | 6.04 |
| Ogun | 23,162 | 3.89 |
| Ekiti | 4,258 | 1.27 |
| Ondo | 16,659 | 3.35 |
| Lagos | 58,750 | 4.59 |

Significantly, the main periods of heightened malaria cases in Oyo, Osun, Ogun, Ekiti, Ondo, and Lagos states appear to be during June and September. When considering all six states, Ondo State recorded the highest levels of rainfall, while Osun State had the lowest temperatures, as evident in Fig 2 and 3.

Given the variability in weather indicators among states, it is more informative to conduct state-based investigations to enhance comprehension of the relationships between malaria incidence, rainfall, and temperature. This approach can help uncover specific patterns and factors contributing to malaria outbreaks in each state individually.

Fig S1–S2 included in the supplementary file also illustrate trends in malaria mortality and diagnosed cases among pregnant women in all states. Though with fewer cases, these diagnosed malaria cases often exhibit patterns similar to those observed in the connections between documented cases of uncomplicated malaria, rainfall, and temperature across different states.

### Autoregressive Integrated Moving Average (ARIMA) models

Following the augmented Dickey-Fuller test to evaluate variance stability, it was confirmed that the variability of malaria cases, along with rainfall and temperature data in Oyo State, remained stable. Subsequently, the ACF and PACF were analysed. For the malaria cases in Oyo State, the auto ARIMA algorithm from the forecast package was used to select a Seasonal ARIMA (SARIMA) (0, 1, 2) (1, 0, 0)12 model. The selection of this model was made based on the criterion of achieving the lowest AIC, determined through a grid search.

Similar model fitting procedures were carried out for malaria caseload data in Osun and Ondo states. In these states, the selected model was SARIMA (1, 0, 0) (0, 0, 0)12. For Ogun state, the SARIMA (1, 1, 1) (0, 1, 1)12 model was identified as the most appropriate for malaria morbidity, having the lowest AIC.

For Ekiti State, the optimal model for malaria caseloads was SARIMA (1, 0, 0) (0, 0, 1)12. Lastly, SARIMA (0, 1, 1) (0, 0, 0)12 was chosen as the best-fitting model for analysing malaria cases in Lagos State.

Although these SARIMA models have limitations in terms of input variables, for the comprehensive planning and evaluation of malaria interventions, they offer valuable insight into the temporal patterns of malaria morbidity in their respective states. These models serve as important tools for forecasting and monitoring trends in malaria cases, aiding in the allocation of resources and the design of targeted interventions to combat malaria in the Southwest region.

As more data becomes available and additional factors are incorporated into the models, they can be further refined to enhance their accuracy and applicability in informing evidence-based strategies for malaria control. Continuous monitoring and evaluation of malaria interventions will remain essential for achieving sustainable progress in malaria elimination efforts across the Southwest region.

### Cross-correlation analysis

The study examined the correlation coefficients among malaria cases, rainfall, and temperature across the Southwest states in Nigeria to comprehend their dynamics and implications for malaria transmission. Detailed results are provided in Table 5, which highlights the varied patterns observed in each state.

**Table 5.** Cross-correlation analysis results of the connection between mean monthly rainfall, temperature and confirmed malaria cases for the Southwest states.

| State | Variable | Lag 0 | Lag 1 | Lag 2 |
| --- | --- | --- | --- | --- |
| Oyo | Rainfall | 0.17 | 0.11 | -0.11 |
|  | Temperature | 0.05 | 0.14 | - |
| Osun | Rainfall | 0.22 | 0.28 | 0.25 |
|  | Temperature | - | 0.06 | 0.04 |
| Ogun | Rainfall | 0.18 | 0.27 | -0.21 |
|  | Temperature | - | 0.17 | 0.29 |
| Ekiti | Rainfall | - | 0.13 | -0.03 |
|  | Temperature | - | -0.10 | 0.08 |
| Ondo | Rainfall | 0.20 | 0.15 | -0.03 |
|  | Temperature | -0.11 | 0.00 | 0.17 |
| Lagos | Rainfall | - | 0.09 | -0.14 |
|  | Temperature | - | 0.01 | 0.02 |

\* Lag 0, Lag 1, Lag 2: Refers to time intervals of 0, 1, and 2 months, indicating the connection between malaria cases, rainfall, and temperature.

These distinct correlation patterns provide insight into the complex interplay between meteorological factors and malaria cases across various Southwest states. In the subsequent cross-correlation analysis, after applying pre-whitening to the series of malaria cases with rainfall, it becomes clear that noteworthy correlations between rainfall and malaria cases consistently manifest with a one-month lag across the Southwest states.

Further analysis of pre-whitened series of malaria cases, rainfall, and temperature data uncovers significant insights into the temporal associations among these variables across different Southwest states. For Osun state, the analysis reveals that the only significant correlations exist at a one-month lag for both rainfall and temperature concerning malaria cases.

Conversely, for Oyo State, Ogun State, Ekiti State, Ondo State, and Lagos State, significant correlations between rainfall and malaria cases, following pre-whitening, appear consistently at a one-month lag. In contrast, temperature exhibits significant correlations at a two-months lag concerning malaria cases in these states.

These results underscore the complex and state-specific dynamics in the relationships between weather factors and malaria caseloads. They emphasize the importance of accounting for temporal lags in comprehending these connections.

Across all states, the analysis indicates that rainfall and increase temperatures precede the increase in malaria cases, providing valuable insights into the timing of malaria incidence.

## Discussion

This study assessed the dynamics of malaria morbidity in Nigeria’s Southwest region, using data from the DHIS2 repository. The outcomes showed distinct patterns of malaria morbidity across the different states in the Southwest zone. These findings corroborate prior investigations studying the relationship between meteorological factors and malaria morbidity countrywide (8,17,18,23). Importantly, however, this study is the first in the region showing the connection between malaria cases and weather variations across the Southwest states at both spatial and temporal dimensions using data from a single source (DHIS2).

A study conducted in certain local government areas in Ondo State revealed (24) that among other factors that result in increased malaria incidence, the most significant of all was temperature. This happens to correlate with the findings in this study, namely that temperature lead malaria incidence by two months. Similarly, our findings also correlates with a study from Ogun State (25) that investigated how climatic factors affected malaria cases in pregnant women and found a link between maximum temperature, humidity, and rainfall with malaria cases. Likewise, our chosen SARIMA model correlates with a model in the study area (25). Findings from a study by Omogunloye et al. (17) also show that maximum temperatures were statistically significant with malaria prevalence in different local government areas in Lagos State, which correlate with findings in this study. Likewise, the findings for the current study have been shown to be similar to those conducted in Jos and Kano states (23), where it was found that the association between malaria incidence and local weather varies at different lag times and direction. Additionally, a study conducted in one local government area (Ado-Ekiti) in Ekiti State found that, increased rainfall and with moderate high temperature would proliferate mosquito species (26). Findings from A. Akinbobola et al. (27) indicated a robust association between climatic variables and malaria incidence, with particular emphasis on maximum temperature and relative humidity in central urban area of Ibadan in Oyo State, which aligns with the findings of the current study.

The increasing seasonal pattern observed across the Southwest states (refer to Fig S3) could stem from the progressive enhancements in the DHIS2 platform for data collection and the consistent rise in malaria case reporting rates. These developments likely contribute to the enhanced reliability of DHIS2 data for predicting malaria morbidity.

While previous studies have assessed the relationship between rainfall and malaria cases within specific localities and communities, employing survey and hospital data (28,29), the present findings using data from the DHIS2 presents a distinct contribution, offering valuable insights. The study underscores the need to recognize as significant a diversity in malaria transmission throughout Southwest Nigeria. From the findings, there is the need for tailored intervention strategies at the state level, rather than adopting a one-size-fits-all approach for the entire zone. This approach may potentially be a more efficient deployment of limited resources, thereby enhancing the effectiveness of interventions.

However, caution needs to be taken when interpreting these findings, due to certain limitations. Firstly, the incidence and meteorological dataset was constrained to monthly observations, potentially overlooking nuanced variations within the data. Furthermore, the model did not encompass other potential malaria risk factors alongside meteorological variables, therein limiting a comprehensive assessment of critical drivers across the southwest states of Nigeria. Factors such as demographics, socioeconomic indicators, public health measures, population mobility, changes in urban planning, geographic characteristics, and non-reporting individuals’ knowledge, attitudes, and behaviours regarding malaria might have influenced the outcomes alongside weather variables. Nevertheless, this research establishes fundamental health data, serving as a cornerstone for future malaria intervention strategies in the Southwest states.

## Conclusion

Using data from the DHIS2 repository spanning from 2013 to 2021, this study has shown diverse malaria patterns within Nigeria’s Southwest zone, with a specific focus on the region’s six constituent states. The investigation shows the correlations between malaria cases and both rainfall and temperature, while considering the combined influence of these meteorological factors across the Southwest states.

This study shows the need for developing targeted and effective malaria interventions across each state of the Southwest regions of Nigeria rather than adopting one a size fit all approach across the zone. By understanding the relationships between malaria incidence and variations in rainfall or temperature, government, healthcare authorities and policymakers can roll out measures tailored to the unique requirements of each state within the region, thereby enhancing disease control initiatives.

Future work includes developing a comprehensive mathematical model that will consider in detail the consequences of various interventions on malaria across the diverse states in Nigeria’s Southwest zone. This mathematical framework will be a helpful tool for simulating and appraising the potential outcomes of diverse control strategies, facilitating evidence-based decision-making in resource allocation and the implementation of measures to effectively reduce the burden of malaria across each state of the southwest, Nigeria.

## Data Availability

The malaria and the meteorological datasets are available on request from the Nigeria NMEP and Nigeria Meteorological Agency (Nimet) respectively.

## Supporting information

**S1 Fig: Monthly Aggregated Malaria in Pregnancy Patterns across Six States (Oyo, Osun, Ogun, Ekiti, Ondo, Lagos) from 2013 to 2021.**

**S2 Fig: Monthly Aggregated Patterns of Malaria Attributable Deaths (<5 years) across Six States (Oyo, Osun, Ogun, Ekiti, Ondo, Lagos) from 2013 to 2021.**

**S3 Fig: Seasonal, Trend, and Residual Analysis of Uncomplicated Malaria Data Series Decomposed by LOESS for Six States (Oyo, Osun, Ogun, Ekiti, Ondo, Lagos) Monthly from 2013 to 2021.**

**Dataset 1: Clinical Data Set Describing Uncomplicated Malaria Cases, Organized by Year, Month, and State.**

## Acknowledgements

We sincerely acknowledge the valuable contributions of the National Research Foundation (NRF) and the University of Cape Town (UCT) postgraduate funding office (PGFO) for their support towards my PhD studies. Additionally, we extend appreciation to the Bill and Melinda Gates Foundation for their generous contributions.

It is important to note that any opinions expressed, and conclusions drawn in this work solely reflect the authors’ own views and do not necessarily represent the positions or perspectives of the NRF and PGFO.

Thanks also goes to Mr. Chukwu Okoronkwo and Mr. Cyril Ademu for their exceptional assistance in data preparation and facilitating the necessary permits for using the data obtained from the National Malaria Elimination Program (NMEP). Their support has been invaluable to the success of this research. Noelle van Biljon also provided valuable insight during the analysis.

## Author Contributions

Conceptualization, O.Y. and S.S.; Data curation, C.O and C.A. Formal analysis, O.Y. Writing - original draft, O.Y. Writing - review & editing, O.Y, C.O, C.A, and S.S. All authors have read through the manuscript and agreed to the published version.

## References

1. World malaria report 2023 [Internet]. [cited 2024 Apr 5]. Available from: https://www.who.int/publications-detail-redirect/9789240086173

2. WHO. World Malaria Report. 2021. Available online: https://www.who.int/news-room/fact-sheets/detail/malaria (accessed on 28 September 2023).

3. Oladipo HJ, Tajudeen YA, Oladunjoye IO, Yusuff SI, Yusuf RO, Oluwaseyi EM, et al. Increasing challenges of malaria control in sub-Saharan Africa: Priorities for public health research and policymakers. Ann Med Surg. 2022 Aug 18;81:104366. 10.1016/j.amsu.2022.104366 PMID: 36046715.

4. WEB_7784 WMR - Nigeria 2022_2408.pdf [Internet]. [cited 2023 Oct 5]. Available from: https://www.afro.who.int/sites/default/files/2023-08/WEB_7784%20WMR%20-%20Nigeria%202022_2408.pdf

5. Sokunbi TO, Omojuyigbe JO, Bakenne HA, Adebisi YA. Nigeria End Malaria Council: What to expect. Ann Med Surg. 2022 Sep 15;82:104690. 10.1016/j.amsu.2022.104690. PMID: 36148088.

6. Blanford JI, Blanford S, Crane RG, Mann ME, Paaijmans KP, Schreiber KV, et al. Implications of temperature variation for malaria parasite development across Africa. Sci Rep. 2013 Feb 18;3:1300. 10.1038/srep01300. PMID: 23419595.

7. Ugwu CLJ, Zewotir T. Evaluating the Effects of Climate and Environmental Factors on Under-5 Children Malaria Spatial Distribution Using Generalized Additive Models (GAMs). J Epidemiol Glob Health. 2020 Dec;10(4):304–314. 10.2991/jegh.k.200814.001. PMID: 33009733.

8. Akinbobola A, Omotosho JB. Predicting malaria occurrence in southwest and north central Nigeria using meteorological parameters. Int J Biometeorol. 2013 Sep;57(5):721–728. 10.1007/s00484-012-0599-6. PMID: 23104425.

9. Silal SP, Barnes KI, Kok G, Mabuza A, Little F. Exploring the seasonality of reported treated malaria cases in Mpumalanga, South Africa. PloS One. 2013;8(10):e76640. 10.1371/journal.pone.0076640. PMID: 24204650.

10. Awine T, Malm K, Peprah NY, Silal SP. Spatio-temporal heterogeneity of malaria morbidity in Ghana: Analysis of routine health facility data. PloS One. 2018;13(1):e0191707.10.1371/journal.pone.0191707. PMID: 29377908.

11. Mukhtar AYA, Munyakazi JB, Ouifki R. Assessing the role of climate factors on malaria transmission dynamics in South Sudan. Math Biosci. 2019 Apr;310:13–23. 10.1016/j.mbs.2019.01.002. PMID: 30711479.

12. Abiodun GJ, Witbooi PJ, Okosun KO, Maharaj R. Exploring the Impact of Climate Variability on Malaria Transmission Using a Dynamic Mosquito-Human Malaria Model. Open Infect Dis J. 2018;10:88–100. 10.2174/1874279301810010088. PMID: 30906484.

13. Craig MH, Kleinschmidt I, Nawn JB, Le Sueur D, Sharp BL. Exploring 30 years of malaria case data in KwaZulu-Natal, South Africa: part I. The impact of climatic factors. Trop Med Int Health TM IH. 2004 Dec;9(12):1247–1257. 10.1111/j.1365-3156.2004.01340.x. PMID: 15598256.

14. Imai C, Cheong HK, Kim H, Honda Y, Eum JH, Kim CT, et al. Associations between malaria and local and global climate variability in five regions in Papua New Guinea. Trop Med Health. 2016;44:23. 10.1186/s41182-016-0021-x. PMID: 27524928.

15. Okunlola OA, Oyeyemi OT. Spatio-temporal analysis of association between incidence of malaria and environmental predictors of malaria transmission in Nigeria. Sci Rep. 2019 Nov 25;9(1):17500. 10.1038/s41598-019-53814-x. PMID: 31767899.

16. Ayanlade A, Sergi C, Ayanlade OS. Malaria and meningitis under climate change: initial assessment of climate information service in Nigeria. Meteorol Appl. 2020;27(5):e1953. 10.1002/met.1953.

17. Omogunloye OG, Abiodun OE, Olunlade OA, Epuh EE, Asikolo I, Odumosu JO. Modeling Malaria Prevalence Rate in Lagos State Using Multivariate Environmental Variations. Geoinformatics FCE CTU. 2018 Aug 23;17(1):61–86. 10.14311/gi.17.1.5.

18. Makinde OS, Abiodun GJ, Ojo OT. Modelling of malaria incidence in Akure, Nigeria: negative binomial approach. GeoJournal. 2021 Jun 1;86(3):1327–1336. 10.1007/s10708-019-10134–x.

19. Segun OE, Shohaimi S, Nallapan M, Lamidi-Sarumoh AA, Salari N. Statistical Modelling of the Effects of Weather Factors on Malaria Occurrence in Abuja, Nigeria - PubMed [Internet]. [cited 2023 Oct 7]. Available from: 10.3390/ijerph17103474. PMID: 32429373

20. An Agricultural Atlas of Nigeria by Agboola, S. A.: Very Good Hardback (1979) 1st. Edition | KENYA BOOKS Books on Kenya & Africa [Internet]. [cited 2023 Jul 26]. Available from: https://www.abebooks.com/first-edition/Agricultural-Atlas-Nigeria-Agboola-S-A/13806116026/bd

21. Constraints to Forest Policy Implementation in the Southwest Nigeria: Causes, Consequences and Cure - PDF Free Download [Internet]. [cited 2023 Oct 8]. Available from: 10.5923/j.re.20120202.06

22. 7.6 Estimation and model selection | Forecasting: Principles and Practice (2nd ed) [Internet]. [cited 2023 Jun 7]. Available from: https://otexts.com/fpp2/estimation-and-model-selection.html

23. Akinbobola A, Hamisu S. Malaria and Climate Variability in Two Northern Stations of Nigeria [Internet]. [cited 2023 Oct 7]. Available from: 10.4236/ajcc.2022.112004

24. Ekpa DE, Salubi EA, Olusola JA, Akintade D. Spatio-temporal analysis of environmental and climatic factors impacts on malaria morbidity in Ondo State, Nigeria.

25. Sadiq B, Brown P. Assessing the Impact of Climatic Variables on Malaria Cases among Pregnant Women in South-Western Nigeria. Univers J Public Health. 2017 Dec 1;5:392–402. 10.13189/ujph.2017.050707.

26. Salau OR. Dependence of Malaria Disease on Changes in Rainfall and Temperature in Ado Local Government Area of Ekiti State, Nigeria. Int J Res. 2016 May 20;3(9):378–87.

27. Akinbobola A, Ikiroma IA. Determining Malaria Hotspot Using Climatic Variables and Geospatial Technique in Central Urban Area of Ibadan, Southwest, Nigeria. J Climatol Weather Forecast [Internet]. 2018 [cited 2023 Oct 13]; Available from: https://www.omicsonline.org/open-access/determining-malaria-hotspot-using-climatic-variables-and-geospatial-technique-in-central-urban-area-of-ibadan-southwest-nigeria-2332-2594-1000225-100031.html

28. Matthew OJ. Investigating climate suitability conditions for malaria transmission and impacts of climate variability on mosquito survival in the humid tropical region: a case study of Obafemi Awolowo University Campus, Ile-Ife, south-western Nigeria. Int J Biometeorol. 2020 Mar;64(3):355–365. 10.1007/s00484-019-01814-x. PMID: 31655868.

29. Awosolu OB, Yahaya ZS, Farah Haziqah MT, Simon-Oke IA, Fakunle C. A cross-sectional study of the prevalence, density, and risk factors associated with malaria transmission in urban communities of Ibadan, Southwestern Nigeria. Heliyon. 2021 Jan;7(1):e05975. 10.1016/j.heliyon.2021.e05975. PMID: 33521357.

